# Comparison of Saliva and Mid-Turbinate Swabs for Detection of COVID-19

**DOI:** 10.1101/2021.12.01.21267147

**Authors:** Jianyu Lai, Jennifer German, Filbert Hong, S.-H. Sheldon Tai, Kathleen M. McPhaul, Donald K. Milton, for the University of Maryland StopCOVID Research Group

## Abstract

**Background:** Saliva is an attractive sample for detecting SARS-CoV-2. However, contradictory reports exist concerning the sensitivity of saliva versus nasal swabs.

**Methods:** We followed close contacts of COVID-19 cases for up to 14 days from last exposure and collected self-reported symptoms, mid-turbinate swabs (MTS), and saliva every two or three days. Ct values, viral load, and frequency of viral detection by MTS and saliva were compared.

**Results:** 58 contacts provided 200 saliva-MTS pairs; 14 contacts (13 with symptoms) had one or more positive samples. Saliva and MTS had similar rates of viral detection (p=0.78) and substantial agreement (κ=0.83). However, sensitivity varied significantly with time since symptom onset. Early on (days -3 to 2), saliva had 12 times (95%CI: 1.2, 130) greater likelihood of viral detection and 3.2 times (95% CI: 2.8, 3.8) higher RNA copy numbers compared to MTS. After day 2 post-symptoms, there was a non-significant trend toward greater sensitivity using MTS.

**Conclusion:** Saliva and MTS demonstrated high agreement making saliva a suitable alternative to MTS for COVID-19 detection. Saliva was more sensitive early in the infection when transmission is most likely to occur, suggesting that it may be a superior and cost-effective screening tool for COVID-19.

## Introduction

The U.S. Centers for Disease Control and Prevention (CDC) recommend the use of upper respiratory specimens, including but not limited to nasopharyngeal, mid-turbinate nasal, anterior nasal and saliva specimens for the initial diagnosis of COVID-19 ^1^. Although nasopharyngeal swabs (NPS) are considered to be the standard for the detection of COVID-19 by most researchers, collection requires the use of trained professionals, can cause discomfort to the patients, and may pose greater risks to healthcare workers during sample collection ^1–4^. Mid-turbinate swabs (MTS) are sometimes used as an alternative to NPS in an effort to reduce patient discomfort and occupational exposures to healthcare workers ^4–6^. Compared to swab-based collection, saliva is even less invasive, more affordable, and can be self-collected with minimal or no supervision ^1,7,8^.

Existing studies focusing on the sensitivity of NPS compared to MTS, and NPS compared to saliva have produced contradictory results ^2,4,9–11^. Few studies directly compare saliva and MTS specimens. Furthermore, there is strong evidence that pre-symptomatic transmission results in higher secondary attack rates for both symptomatic and asymptomatic transmission ^12,13^.

However, most of the existing studies only looked at detection of symptomatic cases after symptom onset ^2,4,9,10^ and few looked at detection sensitivity starting with the pre-symptomatic period. Therefore, research that conducts a direct comparison of MTS and saliva, including an assessment of sensitivity over time (starting during the pre-symptomatic period) is critical to identifying optimally sensitive methods for early detection and effective control of SARS-CoV-2 transmission.

The purpose of this study was to compare the sensitivity of MTS and saliva specimens for detecting SARS-CoV-2 by actively following close contacts of COVID-19 cases and collecting MTS and saliva samples for real-time reverse transcription polymerase chain reaction (RT-PCR) during their post-exposure quarantine period.

## Results

We enrolled 58 individuals with known close contact with an active COVID-19 case. Contacts provided a total of 200 saliva and MTS pairs. The number of days of sample collection per participant ranged from one to seven. Among the contacts, 14 (24%) had at least one positive sample including 11 with both positive saliva and MTS samples over the course of follow-up. One contact had only positive saliva on 3 out of 3 samples (on days -3, 0, and 1 post symptom onset) and 2 had only positive MTS samples; one was positive on 2 of 2 swabs (on days 7 and 10) and one on 1 of 5 swabs (day 21, negative on days 14, 17, 19, and 24). Most of the positive participants (92.9%) were symptomatic, whereas only one (2.3%) participant from the test negative group reported symptoms. Symptomatic participants were enrolled -3 to 14 days since symptom onset and gave samples for up to 24 days from onset of symptoms. Symptoms were mild across the follow-up period. One participant had an oral temperature ≥ 38 °C at the time of sampling; three in total had temperatures ≥ 37.8 °C, and six in total had temperatures ≥ 37.5 °C; all were in the positive group. No other significant differences were identified between the positive and negative groups (Table 1).

**Table 1.** Characteristics of the study population

|  |  | Never positive | Positive for MTS or saliva | All participants |
| --- | --- | --- | --- | --- |
| Number of participants |  | 44 | 14 | 58 |
| Number of sample pairs |  | 159 | 41 | 200 |
| Number of days of sample collection per participant, median (range) |  | 4 (1, 7) | 3 (1, 6) | 3 (1, 7) |
| Female, N (%) |  | 20 (46) | 8 (57) | 28 (48) |
| Age, mean ± SD |  | 26.5 ± 15.5 | 27.3 ± 13.8 | 26.7 ± 15 |
| Age group, N (%) |  |  |  |  |
| <18 |  | 2 (4) | 1 (7) | 3 (5) |
| 18-45 |  | 38 (86) | 11 (79) | 49 (84) |
| >45 |  | 4 (9) | 2 (14) | 6 (10) |
| White, N (%) |  | 30 (68) | 11 (79) | 41 (71) |
| BMI, mean ± SD |  | 25.6 ± 4.9 | 25.2 ± 4.4 | 25.5 ± 4.7 |
| Chronic respiratory illness <sup>a</sup>, N (%) |  | 17 (39) | 5 (36) | 22 (38) |
| Ever smoker, N (%) |  | 1 (2) | 1 (7) | 2 (3) |
| Ever symptomatic <sup>b</sup>, N (%) |  | 1 (2) | 13 (93) | 14 (24) |
| Symptomatic participants | Days since symptom onset at enrollment, median (range) | 2 (-) | 3 (-3, 14 ) | 2.5 (-3, 14 ) |
|  | Overall days since symptom onset of sample collection, median (range) | 6.5 (2, 12) | 5 (-3, 24) | 5 (-3, 24) |
|  | Loss of taste/smell, N (%) | 0 | 2 (15) | 2 (14) |
|  | Median upper <sup>c</sup> respiratory symptoms (IQR) | 3 (1.2, 6.2) | 2 (0, 3) | 2 (0.2, 3) |
|  | Median lower respiratory symptoms (IQR) | 0 (0, 0) | 0 (0, 1) | 0 (0, 1) |
|  | Median systemic symptoms (IQR) | 0.5 (0, 1.8) | 0 (0, 1.2) | 0 (0, 1.8) |
|  | Median gastrointestinal symptoms (IQR) | 0 (0, 0) | 0 (0, 0) | 0 (0, 0) |
|  | Temperature, mean Celsius ± SD | 37 ± 0.3 | 37.2 ± 0.5 | 37.2 ± 0.4 |

<sup>a</sup> Chronic respiratory illness = volunteers with any Chronic obstructive pulmonary disease, Asthma, Other lung diseases.
<sup>b</sup> Group comparison, $p < 0.05$ .
<sup>c</sup> Symptoms at the time of each sample collection visit. Sixteen individual symptoms were rated from 0 to 3. Systemic (max score of 12) = malaise + headache + muscle/joint ache + sweats/fever/chills; Gastrointestinal (max score of 12) = loss of appetite + nausea + vomit + diarrhea; Lower Respiratory (max score of 9) = chest tightness + shortness of breath + cough; Upper Respiratory (max score of 15) = runny nose + stuffy nose + sneeze + earache + sore throat.

### Viral RNA detection in and agreement between saliva and MTS

Among 200 pairs of saliva and MTS samples we detected viral RNA in 32 (16%) of the saliva and 29 (14.5%) of the MTS samples. The frequency of detection was similar for both sample types (p=0.781) (see Supplementary Table S1 online). Cohen’s Kappa demonstrated substantial agreement (κ=0.83) with 26 (14%) positive and 165 (82.5%) negative sample pairs (Table 2). The 14 participants who became positive by either sample type during the follow-up period provided 41 saliva-MTS sample pairs, among which 71% of MTS and 78% of saliva samples were positive (see Supplementary Table S2 online), without respect to time since symptom onset. When focusing on positive participants, however, the agreement was weak (κ=0.43 for all and κ=0.42 for those who were symptomatic) (see Supplementary Table S3a and Table S3b online).

**Table 2.** Viral RNA detection in paired saliva and MTS samples from all participants (N=58) ^a^

| *Saliva Positive* | *MTS Positive* |  | ***Total*** |
| --- | --- | --- | --- |
|  | No | Yes |  |
| No | 165 | 3 | 168 |
| Yes | 6 | 26 | 32 |
| ***Total*** | 171 | 29 | 200 |

<sup>a</sup> Cohen's Kappa between the two sample types was calculated as $\kappa=0.83$

### Comparison of Ct (cycle threshold) values between saliva and MTS

Each RT-PCR reaction contained 10 µL of heat-treated saliva sample or RNA extracted from MTS. Assuming no loss in the process, each reaction represented 7.78 µL of saliva sample or 40 µL of the MTS eluate. The Ct values for paired samples were highly correlated (rho = 0.84, r2 = 0.74, Figure 1a). The Ct values for saliva were on average slightly but significantly greater than for MTS samples (mean difference = 0.64, p=0.01) among all 58 participants (see Figure 1b and Supplementary Fig. S1 online), partially reflecting the difference in their input amounts.

**Figure 1.**
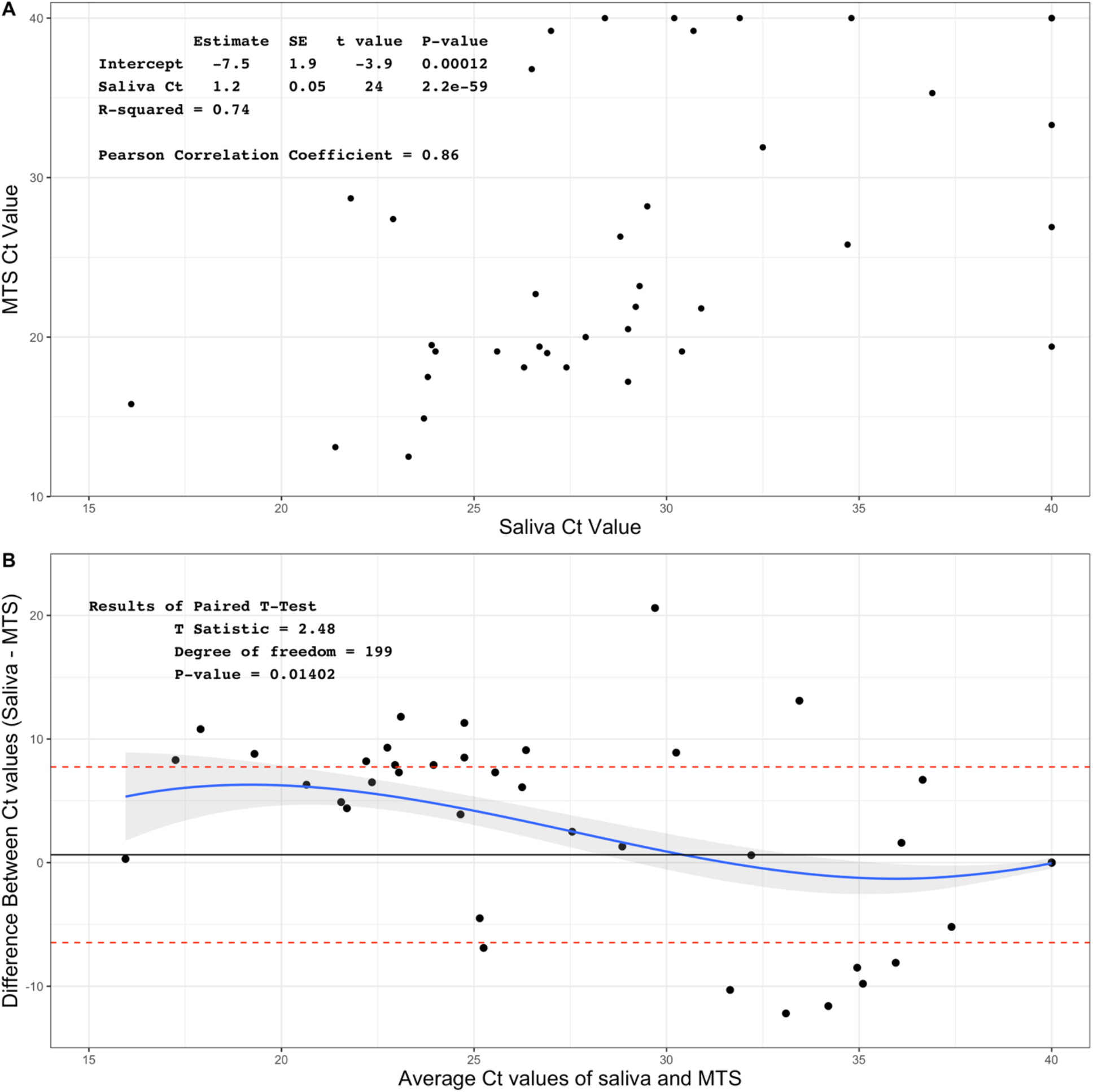
Association between Ct values of saliva and MTS samples. Data were from 58 participants and 400 samples. A. Scatter plot of Ct values of saliva and MTS B. Bland-Altman plot for comparison of saliva and MTS

### Relationship between days since symptom onset, probability of detection, and viral RNA copy numbers

The Ct values among the positive symptomatic participants increased over time (days -3 through 24), along with decreasing viral RNA copy numbers. Saliva tended to have lower Ct values and higher viral RNA copy numbers compared to MTS from days -3 to 1.5, whereas MTS samples had lower Ct values and higher viral load thereafter (Figure 2A and Figure 2B).

**Figure 2.**
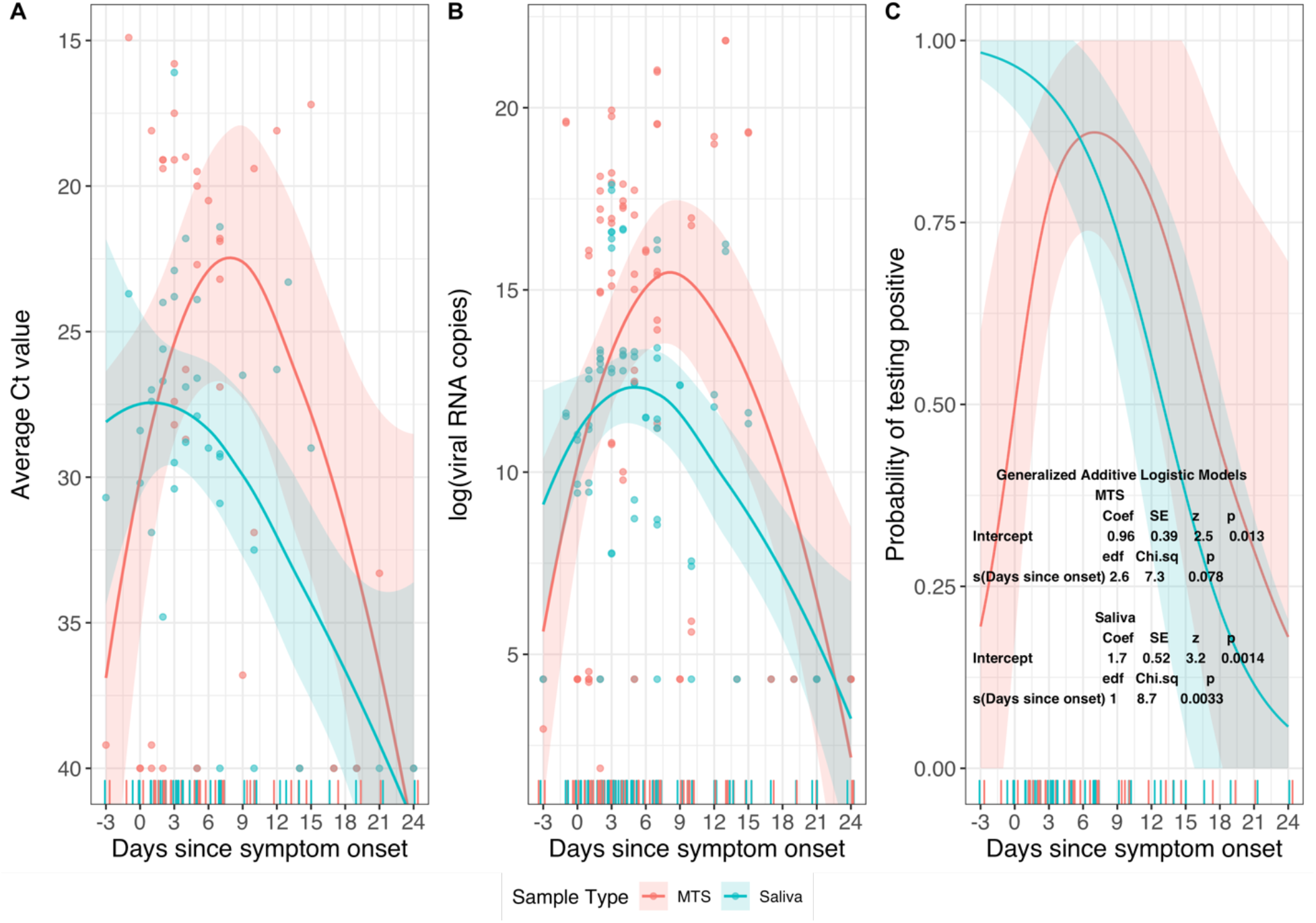
The change of Ct values and probability of testing positive by days since symptom onset. Data were for MTS and saliva samples from 13 participants who provided a total of 40 pairs of samples and had one or more samples positive for SARS-CoV-2 RNA. A. Change of Ct values by days since symptom onset, B. Change of viral RNA copy numbers (natural log scale) by days since symptom onset, C. Probability of being tested positive by days since symptom onset estimated from a generalized additive logistic model.

Among symptomatic participants who had one or more positive saliva or MTS samples, the probability (sensitivity) of detecting viral RNA in saliva samples was 91% (10/11) from day -3 to day 2 (Table 3), was 89% (16/18) from day 3 through 8, and declined significantly thereafter (see Figure 2C and Supplementary Fig. S2 online). The probability of detecting virus in MTS samples from day -3 through day 2 was 45% (5/11), was 94% (17/18) from day 3 through 8, and then declined.

**Table 3.**
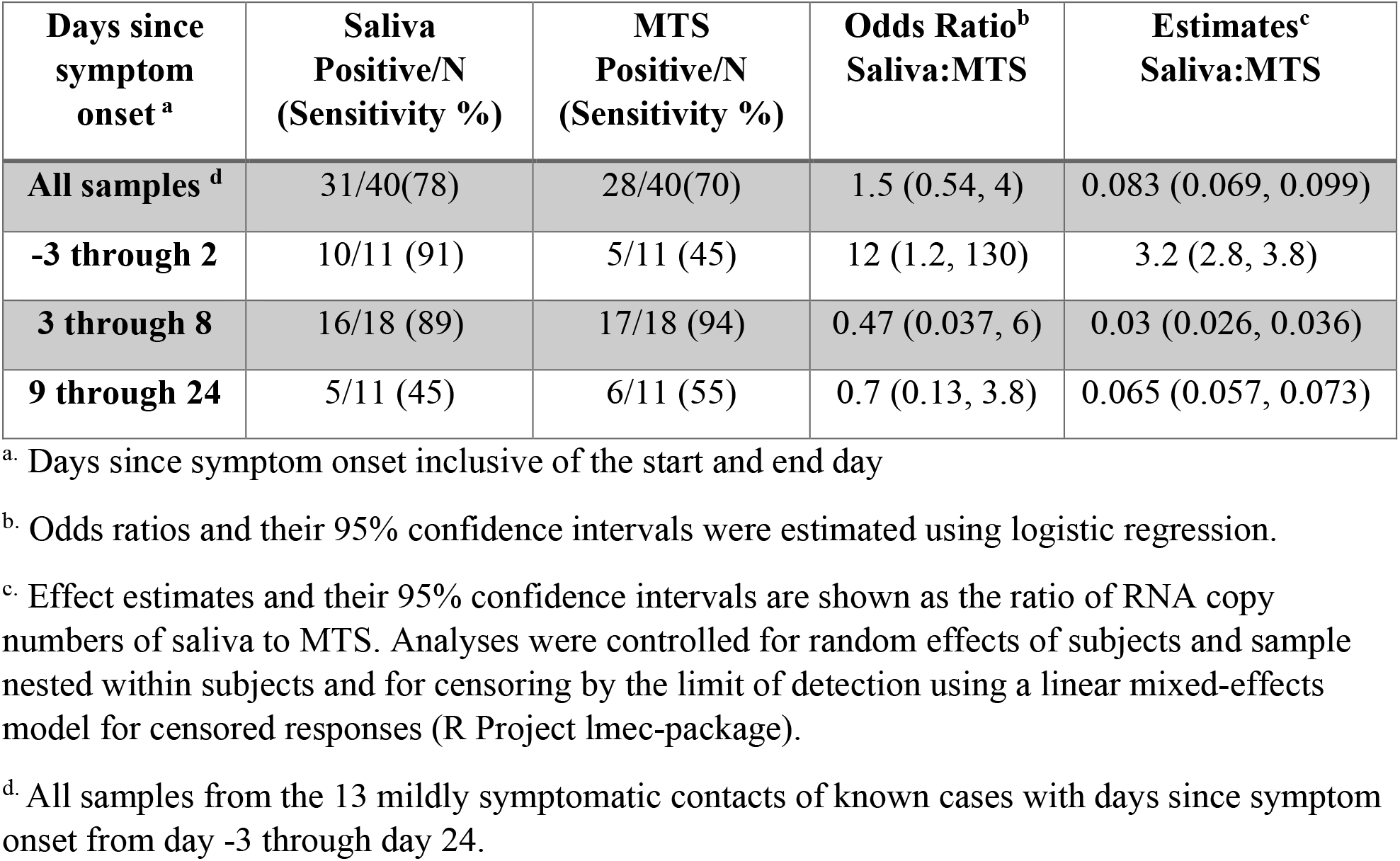
Sensitivity of saliva and MTS and relative odds of detection and ratio of viral RNA copy numbers of SARS-CoV-2 in saliva by day since symptom onset among 13 mildly symptomatic contacts of known cases

Early in the course of infection (days -3 through 2) saliva had 12 times the odds of being positive (95% CI: 1.2, 130) and 3.2 times higher viral RNA copy numbers (95% CI: 2.8, 3.8) compared to MTS. There was a trend toward greater sensitivity and higher viral RNA copy numbers in MTS than saliva samples after day 2 post onset of symptoms (Table 3).

### Asymptomatic case

Only one participant from our study population was an asymptomatic case. They provided one pair of saliva and MTS samples, both of which were positive, with an average Ct value of 25.8 for MTS and 34.7 for saliva (see Supplementary Table S4 online).

## Discussion

Early in the course of infection, saliva was significantly more sensitive than mid-turbinate nasal swabs (MTS). We found that the optimal performance of saliva was in the pre-symptomatic period and was more sensitive than MTS before symptom onset. Several studies have shown that pre-symptomatic transmission plays a more important role than symptomatic and asymptomatic transmission in the spread of SARS-CoV-2 ^12,13^. Furthermore, saliva tended to have lower Ct values and higher viral load compared to MTS from the pre-symptomatic period through the first days post symptom onset. Together, these findings suggest that saliva may be the most effective method for detecting SARS-CoV-2 early during the course of infection.

The CDC and the Infectious Disease Society of American recommendations for COVID-19 testing allow MTS, NPS, oral swabs, anterior nasal swabs, and saliva swabs as well as saliva.^1,14^ Some studies have shown differences in the sensitivity between NPS and MTS. In older, more acutely ill populations, NPS appears to be more sensitive than MTS, especially later in the course of illness (greater than 7 days) ^4,9^. In a study of ambulatory and symptomatic participants whose ages were more evenly distributed, NPS and MTS swabs were highly correlated with a mean of 7 days since onset of symptoms ^15^. Congrave-Wilson et al., in agreement with the current study, found that saliva had the highest sensitivity in the first seven days post COVID-19 onset when using NPS as the reference ^2^. Similarly, Savela et al. noticed that although the peak viral load of SARS-CoV-2 in nasal swabs were higher than saliva, the latter was more likely to be positive in the first six days since the participants’ first positive sample was detected^16^. Becker et al. compared the sensitivity of saliva and NPS for detecting COVID-19 in a convalescent cohort 8-56 days since first symptom and found that NPS performed better ^17^. They also showed that saliva was about 30% less sensitive than NPS in a separate diagnostic cohort, however, days since symptom onset were not reported, so we cannot make direct comparison with our findings. Finally, a systematic review by Bastos et al. found that saliva had similar sensitivity to NPS and costs less ^11^.

Our findings have implications for improving public acceptance of COVID-19 testing, reducing the cost of mass COVID-19 screening, and improving the safety of healthcare workers who conduct testing. These findings are extremely important when considering large-scale screening of COVID-19 in schools and workplaces. In addition to its higher sensitivity in the early stage of the disease as demonstrated in our data, saliva has quite a few other advantages that make it an appealing screening tool. Saliva collection is less invasive and more acceptable to the general population ^7,18^. One of the barriers hindering COVID-19 testing is people’s fear of nasal swabs due to misinformation ^19^. Also, the discomfort brought by nasal swabs may also reduce people’s willingness to get tested regularly, especially among children ^20,21^. With the use of saliva, screening large groups with increased frequency may be more practicable. Saliva is cheaper than swab-based methods, especially if pooled samples are used ^11,22^. Bastos et al. estimated that when sampling 100,000 individuals, using saliva saved more than $600,000 in comparison to using NPS ^11^, using a method that is more expensive than the SalivaDirect method used here. These cost savings are especially important in the context of low resource settings.

Saliva collection is also safer for healthcare workers (HCWs). Amid the pandemic, one of the key concerns among HCWs is the occupational exposure to SARS-CoV-2 aerosols during some medical procedures ^23^. The collection of nasal swabs introduces such exposure via the close interaction between patients and HCWs and by patients’ coughing and sneezing as a result of the procedures ^24^. In contrast, saliva is the only upper respiratory specimen suggested by the CDC that can be self-collected without supervision ^1^ and hence protects HCWs from directly contacting the patients when the samples are being collected. Given all these advantages of saliva compared to NPS, our findings further support the use of saliva for large-scale screening, especially of pre-symptomatic patients.

The current study has several limitations. The contacts enrolled in this study who eventually tested positive for SARS-CoV-2 developed mild, and in some cases, transient infections. Some of these mild (low viral titer) infections would not have been detected by less frequent testing protocols and may not have posed a risk for onward transmission. The sample size of those who tested positive is relatively small. Only one asymptomatic case was identified in our study so we could not compare the sensitivity of the two types of samples among asymptomatic COVID-19 cases. The evidence for using saliva to detect asymptomatic cases was mixed in previous studies ^2,10^ and further studies are needed to clarify this issue.

Future research should focus on the development of rapid saliva tests with high sensitivity and specificity. Tng et al. proposed an amplified parallel antigen rapid test (AP-ART) using saliva to test SARS-CoV-2 with a turnaround time of only 30 minutes^25^. This test was reported to have a sensitivity as high as 97%. However, the researchers did not compare this AP-ART with saliva-based RT-PCR but instead NPS-based RT-PCR and estimated the specificity of this test to be only 90%. As our study showed, this is likely an underestimate of specificity because saliva can be more sensitive than nasal swabs early in the course of the infection. Hence, further studies are needed to evaluate saliva rapid tests with a reference method that is also based on saliva.

In conclusion, the use of saliva is preferable for testing pre-symptomatic populations. It is more acceptable to people, which reduces barriers to testing. It is also more cost effective for individuals to collect their own saliva rather than using highly trained professionals to collect NPS and/or MTS. Finally, self-collected saliva samples eliminate the exposure to aerosols produced by sneezing, coughing and gagging of patients undergoing NPS/MTS.

## Methods

### Study population

We analyzed MTS and saliva sample data from individuals who reported close contact with confirmed COVID-19 cases as part of the University of Maryland StopCOVID study ^26^ from May 2020 to April 2021.

### Questionnaire and sample collection

Participants were followed every two or three days for up to 14 days from their last exposure or until SARS-CoV-2 was detected in their samples. If one or more of their screening samples became positive, results were confirmed by an appropriate clinical diagnostic test and they were recruited to participate in the exhaled breath aspect of the study that also involved the collection of saliva and MTS ^26^. On each day of sample collection, participants answered an online questionnaire to update their current symptoms and medications. For those who reported having any symptom, they also reported their symptom onset date (i.e., “When did you begin to feel sick?”).

The symptoms checklist in the baseline and follow-up questionnaires, as previously described ^26^, included runny nose, stuffy nose, sneezing, sore throat, earache, malaise, headache, muscle and/or joint ache, sweat/feverish/chills, nausea, loss of appetite, vomiting, abdominal pain or diarrhea, chest tightness, shortness of breath, and cough. Participants self-reported for each of these 16 symptoms on a scale of 0 to 3 (0 = “no symptoms,” 1 = “just noticeable,” 2 = “clearly bothersome from time to time, but didn’t stop me from participating in activities,” 3 = “quite bothersome most or all of the time and stopped me from participating in activities”).

For saliva collection, participants were instructed to not eat or drink 30 minutes prior to the visit and then collect approximately 0.5-1 mL of saliva drooled into a plastic collection tube. For MTS collection, trained clinical staff inserted a mid-turbinate swab approximately 1.5-2 inches into one of the participants’ nostrils, rotated once, and then withdrawn. This procedure was repeated in the other nostril for a total of two MTS per participant per visit.

### Laboratory analyses

Saliva samples were processed using the SalivaDirect method ^8^ as previously described ^26^. Briefly, 50 µL of individual saliva samples were treated with Proteinase K (New England Biolabs), heated at 95°C for 5 minutes, and kept at 4°C. MTS from both nostrils were combined and processed as previously described ^26^. Briefly, total nucleic acid was extracted from 200 µL of MTS with MagMax Pathogen RNA/DNA Kit (Applied Biosystems) on KingFisher Duo Prime (Thermo Fisher Scientific), following the manufacturers’ protocols. The sample was eluted in 50 µL of Elution Buffer and kept at 4 °C. MS2 phage was spiked in each heat-treated saliva sample and extraction to control for extraction and PCR failure. RT-PCR was set up on the same day; each reaction consisted of 1X TaqPath 1-Step Master Mix, No ROX, 1X TaqPath COVID-19 Real Time PCR Assay Multiplex (both from Thermo Fisher Scientific), and 10 µL of heat-treated saliva or eluted nucleic acids. Each PCR plate contained a positive control provided in the TaqPath COVID-19 Combo Kit (Thermo Fisher Scientific) and a no template control. Viral loads in saliva and MTS were quantified as previously described^26^. RNA copy numbers were reported per mL for saliva and per sample for MTS. The limit of detection was 75 copies per sample and the limit of quantification was 250 copies per sample. A positive sample was defined as having Ct values < 40 for at least two out of three SARS-CoV-2 targets (ORF1ab, N gene, and S gene)^27^. The average Ct values of all positive targets were used in the following analyses.

### *Statistical* analyses

We analyzed only paired same-day saliva and MTS samples to ensure the comparability of the two samples. Group comparisons were made between participants having a positive result for either sample and those with both samples being negative. Continuous variables (age and BMI) were compared using t-test, and categorical variables were compared using Chi-square test (sex and chronic respiratory illness) and Fisher’s exact test (age group, race, and ever smoker).

To compare the Ct values from saliva and MTS, we conducted paired t-test and Bland-Altman analysis, and calculated the coefficient of determination (i.e., R squared from linear regression) and Pearson correlation coefficient. The Chi-square test was used to explore the relationship between detection and sample types. Cohen’s Kappa was calculated to demonstrate the degree of agreement between the two sample types.

For participants with a positive saliva or MTS sample, we used a generalized additive logistic model ^28^ to estimate and plot the probability of having a positive result by days since symptom onset for the two sample types. We also created a plot using the LOESS (locally weighted smoothing) method with 95% confidence interval for the change of Ct values and viral RNA copy numbers by days since symptom onset for the two sample types. Logistic regression was used to estimate the relative odds of detection of SARS-CoV-2 in saliva over specified intervals since symptom onset.

For the estimate of geometric means of viral RNA copy numbers and the ratio of RNA copy numbers of saliva to MTS, we applied linear mixed-effect models with censored responses^29,30^ to handle censored observations below the limit of detection and control for random effects of subjects and sample nested within subjects.

All the analyses were carried out using RStudio and R (version 4.0.4) ^31^.

### Ethics statement

This study was approved by the University of Maryland Institutional Review Board and the Human Research Protection Office of the Department of the Navy. Electronically signed informed consent was obtained from all participants and questionnaire data were collected and stored with REDCap ^32^.

## Data Availability

The datasets generated during and/or analyzed during the current study were deposited at Open Science Framework (OSF) repository: DOI 10.17605/OSF.IO/9YP3Z (https://osf.io/9yp3z/)

## Acknowledgements

We thank all the other members of the University of Maryland StopCOVID Research Group for their efforts in recruiting participants and sample collection and processing: Oluwasanmi Oladapo Adenaiye, Barbara Albert, P. Jacob Bueno de Mesquita, Yi Esparza, Aaron Kassman, Michael Lutchenkov, Dewansh Rastogi, Maria Schanz, Isabel Sierra Maldonado, Aditya Srikakulapu, Delwin Suraj, Faith Touré, Rhonda Washington-Lewis, Somayeh Youssefi, Stuart Weston, Matthew Frieman, Mara Cai, Ashok Agrawala. We also thank Dr. Jamal Fadul and his clinic in College Park, Maryland, for assistance in recruiting study participants.

## Author Contributions

Conceptualization, D.M., J.G., F.H., S.T., J.L.; Data curation, F.H.; Formal analysis, J.L.; Investigation, S.T., J.G., J.L., K.M.; Methodology, D.M., J.G., S.T., J.L.; Project administration, D.M., F.H.; Writing-original draft, J. L., J.G.; Writing-review & editing, all authors; Supervision, D.M.

