## Supplemental Tables and Figures for "Comparison of Saliva and Mid-Turbinate Swabs for Detection of COVID-19"

Table S1. Association between detection frequency and sample types (MTS vs. saliva) [a]

| *Sample Positive* | *Sample Type* | | ***Total*** |
|---|---|---|---|
| | MTS | Saliva | |
| No | 171 | 168 | 339 |
| Yes | 29 | 32 | 61 |
| ***Total*** | 200 | 200 | 400 |

[a] $\chi^2$=0.077, df=1, p=0.781

Table S2. MTS and saliva for positive participants (N=14)

| | Number of subjects | Number of samples | Number of positive samples, N (%) | Ct value of positive samples, median (range)[a] | GM (95% CI)[b] of all samples | GM (95% CI)[c] of positive samples |
|---|---|---|---|---|---|---|
| MTS | 13 | 41 | 29 (71) | 20 (12, 33) | 2.2 x$10^5$ (1.0 x$10^5$, 4.9 x$10^5$) | 4.5 x$10^6$ (3.5 x$10^6$, 5.9 x$10^6$) |
| Saliva | 12 | 41 | 32 (78) | 27 (16, 35) | 2.3 x$10^3$ (1.2 x$10^3$, 4.3 x$10^3$) | 4.5 x$10^5$ (4.0 x$10^5$, 5.0 x$10^5$) |

[a]. A positive sample was defined as having Ct < 40 in PCR detection for at least two out of three SARS-CoV-2 genes (N, S, ORF1ab)

[b]. GM = geometric mean. The GMs were computed, accounting for samples below the LOD, using a linear mixed-effects model for censored responses (R Project LMEC package) using data for all samples of each sample type with nested random effects of samples within study participant.

[c]. GM were calculated using data from only positive samples of each sample type with nested random effects of samples within study participant.

Table S3a. Viral RNA detection in paired saliva and MTS samples from participants with one or more positive samples (N=14) [a]

| *Saliva Positive* | *MTS Positive* | | ***Total*** |
|---|---|---|---|
| | No | Yes | |
| No | 6 | 3 | 9 |
| Yes | 6 | 26 | 32 |
| ***Total*** | 12 | 29 | 41 |

[a] Cohen's Kappa between the two sample types was calculated as κ=0.43

Table S3b. Viral RNA detection in paired saliva and MTS samples from symptomatic participants with one or more positive samples (N=13) [a]

| *Saliva Positive* | *MTS Positive* | | ***Total*** |
|---|---|---|---|
| | No | Yes | |
| No | 6 | 3 | 9 |
| Yes | 6 | 25 | 31 |
| ***Total*** | 12 | 28 | 40 |

[a] Cohen's Kappa between the two sample types was calculated as κ=0.42

Table S4. Saliva and MTS testing results for the asymptomatic positive participant (N=1)

| | **Ngene Ct** | **Orf1ab Ct** | **Sgene Ct** | **Average Ct** |
|---|---|---|---|---|
| **MTS** | 26.7 | 25.2 | 25.6 | 25.8 |
| **Saliva** | 33.4 | 36.3 | 34.5 | 34.7 |

### Supplemental Figures

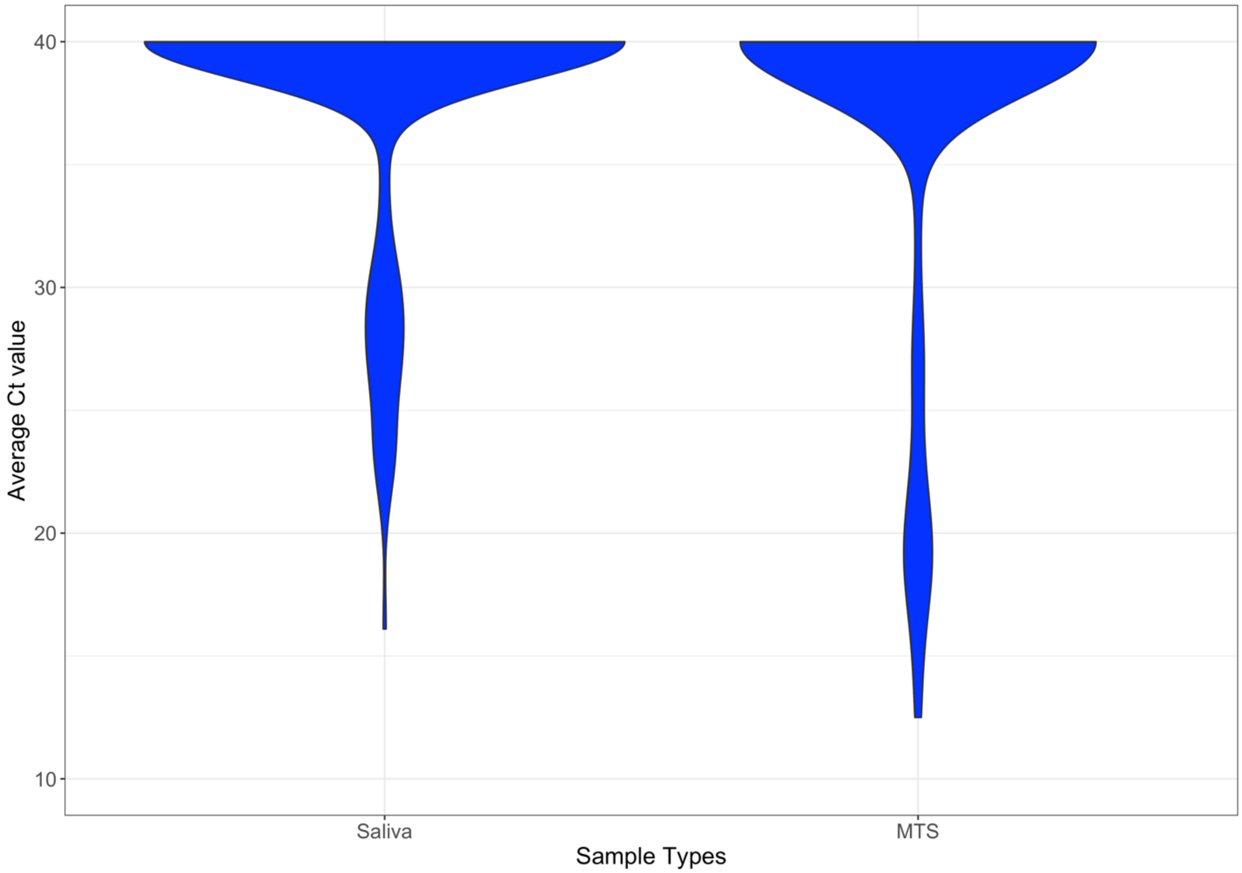


Figure S1. Distribution of Ct values by sample types among 58 participants and 400 samples.

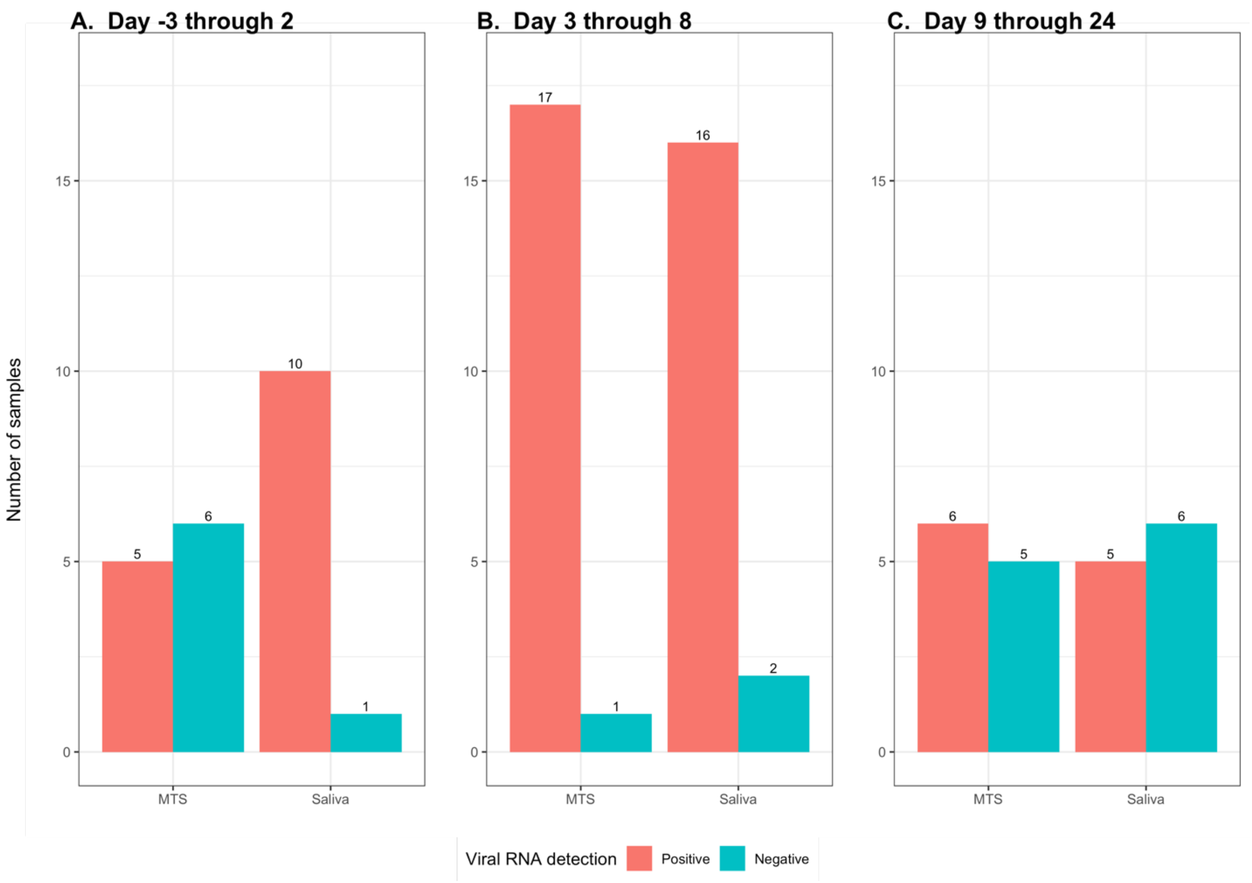


Figure S2. Distribution of samples from 13 symptomatic positive cases by days since symptom onset. A) Days -3 through 2, B) Days 3 through 8, C) Days 8 through 24.
